# Adherence to the TRIPOD statement of clinical prediction models for spinal pain or osteoarthritis: a protocol for a meta-research study

**DOI:** 10.1101/2023.04.19.23288819

**Authors:** Daniel Feller, Roel Wingbermuhle, Tiziano Innocenti, Raymond Ostelo, Alessandro Chiarotto

## Abstract

**Background:** Studies that develop and/or validate a clinical (diagnostic or prognostic) prediction model should be reported with sufficient details to enable the clinician to use the model in clinical practice. Also, an adequate reporting enables researchers to externally validate the model and assess its validity and reliability. To improve the reporting of multivariable prediction models, the TRIPOD checklist was released in 2015.

**Objective:** Evaluate the quality of reporting of prediction model studies conducted in patients with back pain, neck pain or osteoarthritis.

**Methods:** We will perform a systematic review of the literature, searching MEDLINE (via Ovid), Embase, Web of Science, and CINHAL (via EBSCO) for studies that describe a model development, external validation, update, or a combination of these in patients with back pain, neck pain or osteoarthritis. The completeness of the reporting will be evaluated using a specific adherence assessment form of the TRIPOD. Two authors will independently perform the study selection and data extraction processes. A descriptive analysis of the total score and the individual items of the TRIPOD will be produced.

**Ethics and dissemination:** Upon the completion of the study, a manuscript with the results will be submitted for publication in a peer-reviewed journal.

## BACKGROUND

The *“Global Burden of Disease”* study highlights that musculoskeletal (MSK) disorders are among the causes of the highest disability-adjusted life years (DALYs) worldwide. ^1^ Also, MSK disorders are the conditions requiring more rehabilitation in all the categories of age, irrespective of the location. ^1^ Among MSK conditions, back pain, neck pain, and osteoarthritis (OA) are the most burdensome. For example, between 25 and 49 years old, low back pain is ranked number 4 on the list of the disease with the highest DALYs (only after road injuries, HIV/AIDS, and ischaemic heart disease), and OA is number 18 in this ranking between 50 and 74 years old. ^2^ All these data suggest the need for more research on MSK disorders. One field of extensive research relating to MSK conditions is predictive modelling, both for prognostic and diagnostic purposes. Indeed, in recent years we have assisted to an increasing number of prediction models developed for MSK disorders, particularly for spinal conditions and OA. ^3–5^

A prediction model is a statistical model specifically designed to predict the prognosis of a health condition (i.e., prognostic prediction model) or to estimate the probability that a specific disease is present (i.e., diagnostic prediction model). ^6,7^ The development of both prognostic and diagnostic prediction models is executed using statistical methods and machine learning techniques, and they are typically based on a combination of patient characteristics, clinical variables, and laboratory or imaging data. ^7^ Prediction model studies are categorized into model development, model validation, and clinical impact studies. ^8^ Model development studies aim to create a predictive model by selecting pertinent predictors and aggregating them statistically into a multivariable model. On the other hand, external validation studies aim to evaluate the model’s performance on a new and independent dataset. The purpose of a validation study is to assess the generalizability of the model and to ensure that it performs accurately and reliably in different populations or settings with similar inclusion and exclusion criteria. ^6,7^

Clinicians can use predictive models to inform decision-making, such as determining the best course of treatment for a patient based on their individual prognosis. However, this can be achieved only if the development or validation study is described in sufficient detail to enable the clinician to use the model in clinical practice. ^8–10^ Also, an accurate reporting of the model’s development or validation study allows other researchers to assess the validity and reliability of the model, replicate the study if necessary, and build upon the findings in future research. ^10^

To improve the reporting of studies that develop, validate, or present prediction models, in 2015, an international group of experts in the field of prediction modelling published the TRIPOD (*“Transparent Reporting of a multivariable prediction model for Individual Prognosis or Diagnosis”*) statement. ^8^ The TRIPOD is a guideline comprising 22 items that should be included in reporting a study involving a prediction model. The checklist covers various aspects of the study, including the rationale and objectives of the model, the study design and population, the statistical methods used, and the performance measures of the model. ^8,11^

Although the reporting of prediction model studies is crucial and guidelines such as TRIPOD are available, studies in several biomedical fields (e.g., oncology, neurology) indicate that the quality of reporting of prediction models is insufficient. ^12–14^ To ensure uniformity in measuring adherence to the TRIPOD statement, in 2019 Heus et al. published an adherence scoring system. The adherence assessment form (AAF) created contains all the main items of the original TRIPOD, of which ten are divided into sub-items. Most items are then split into multiple adherence elements to take into account that the original TRIPOD items often require multiple elements to be reported. Every adherence element of the AAF are statements with four possible answers (i.e. yes, no, referenced, and not applicable) that collectively can be used to determine the singles items and overall adherence to the TRIPOD. ^11^

Currently, no studies have assessed the completeness of the reporting in studies that develop or validate a prediction model for patients with back pain, neck pain or OA. Therefore, the objective of the present study is to evaluate the quality of reporting of prediction model studies conducted in patients with back pain, neck pain, or OA.

## MATERIAL AND METHODS

We will report the results of this study using an adapted version of the *“Preferred Reporting Items for Systematic Reviews and Meta-analyses”* (PRISMA) checklist for meta–research studies ^15^ or, if it will be available at the time of the reporting, using the “MethodologIcal STudy reporting Checklist” (MISTIC). ^16^

### Inclusion criteria

We will include primary studies that developed and/or externally validated a multivariable prediction model in patients (without age restriction) experiencing back pain, neck pain, or OA, irrespective of study design and outcome measurement. In this context, we define a multivariable prediction model as any model that combines two or more predictors to estimate the probability that a specific disease is present (i.e., diagnostic model) or to predict the prognosis of a health condition (i.e., prognostic model). ^7^ Also, we will include studies that aim to discover the incremental value of adding one or more predictors to an existing model. ^11^

We will only include studies published between the 1^st^ of January 2016 and the 16^th^ of March 2023 in English, Italian, or Dutch. We will use this time restriction because the TRIPOD checklist was published in 2015 and we assume some delay from publication and possible widespread use; meanwhile, we choose to restrict the inclusion to the languages mentioned above because those are the mother languages of the authors.

### Exclusion criteria

We will exclude secondary research (e.g., systematic reviews), editorials, conference abstracts, non-peer reviewed articles, prognostic factor studies ^17^, studies primarily interested in determining a disorder’s aetiology, studies examining the clinical impact of a clinical prediction model, and predictive models that used genetic data. We will also exclude studies focusing on screening tools for risk prediction (e.g., STarT Back Tool, Orebro Musculoskeletal Pain Questionnaire).

### Study selection process

Primary studies will be searched in biomedical databases, through forward and backward citation tracking strategies (Web of Science), and using the bibliographic information of relevant reviews found in the selection process that aimed to summarize the evidence regarding predictive models in back pain, neck pain and OA.

The biomedical databases investigated will be MEDLINE (via Ovid), Embase, Web of Science, and CINHAL (via EBSCO).

Supplementary material 1 reports the search strategy used in all the databases.

Two researchers will independently perform the study selection process, firstly by title/abstract and finally by full text. Any disagreement will be resolved by consensus or by the decision of a third author.

We will use the online electronic systematic review software package (Rayyan QCRI) to organize and track the selection process. ^18^

### Data extraction

The data extraction process will be conducted independently by two reviewers. Any discrepancies will be resolved with a consensus between the two authors and eventually by a third author’s decision.

We will extract the following data:

- First author and year of publication
- Study design (e.g., prospective cohort study)
- Objective (model development and/or validation, update of a model with one or more predictors)
- Type of prediction model (prognostic or diagnostic)
- Type of statistical/machine learning methods used (e.g., logistic regression, random forest)
- Outcome(s) predicted (e.g., pain intensity)
- Declared use of the TRIPOD as reporting guideline (yes/no)
- Adherence to the TRIPOD guideline

The adherence to the TRIPOD guideline will be collected using the specific AAF developed in 2018 by Heus et al. ^11^ Depending on whether a report describes model development, external validation, a combination of these, or the incremental value of adding one or more predictors, we will use the adequate AAF. ^11^ If a study contains multiple prediction models developed or validated, we will base the scoring on the primary model as stated by the authors or, if this is not stated, on the first model reported in the methods section. ^11^

The AAF will be piloted among the reviewers responsible for the data extraction process using a randomly selected 5% of all the included studies. The piloting results will be discussed to reach a consensus about the data extraction phase.

### Data synthesis

The data collected will be summarized descriptively with the aid of tables and graphs. First, we will compute the adherence to every single TRIPOD item. If all adherence elements of a particular item are answered with a “yes”, “not applicable”, or “referenced”, we will score that TRIPOD item as “1”. If all the adherence elements are answered as “not applicable”, we will score the item as “not applicable” overall. Otherwise, that item will be scored as a “0” (non-adherence). ^11^ The adherence to the single TRIPOD items will then be calculated by dividing the number of studies adhered to that specific item by the number of studies in which the item was applicable.

We will calculate the overall TRIPOD adherence score by dividing the sum of the adhered TRIPOD items by the total number of applicable items for that report. The overall score will be presented as a percentage of adherence. To summarize the results, we will report the median with the interquartile range for the overall score. ^11^ In addition, as a subgroup analysis, we will descriptively compare the overall TRIPOD adherence score between study type (development, validation, development and validation, update by adding one or more predictors), type of disorder (back pain, neck pain and OA), type of prediction model (prognosis and diagnosis), and year of publication.

The statistical analysis will be carried out using R and RStudio. ^19^

## Data Availability

All data produced in the present study are available upon reasonable request to the authors

## ETHICS AND DISSEMINATION

An adequate reporting of a clinical prediction model study is important for many reasons. For example, it enables the clinician to implement the model directly in clinical practice, permits the external validation of the model, and allows researchers to assess the validity and reliability of the model. To the best of our knowledge, no study has ever investigated the completeness of the reporting in studies that develop or validate a prediction model for patients with back pain, neck pain, and OA. Therefore, this study will provide valuable information that may improve future research in the development and/or validation of prediction models. After completion of the study, a manuscript with results will be submitted for publication in a peer-reviewed journal.

## AUTHOR CONTRIBUTIONS

DF and AC conceived and designed the study protocol. All the authors provided input and approved the final version of the protocol.

## SUPPLEMENTARY MATERIAL 1

| Database searched | Platform | Years of coverage | Records | Records after duplicates removed |
| --- | --- | --- | --- | --- |
| Medline ALL | Ovid | 1946 - Present | 1707 | 1702 |
| Embase | Embase.com | 1971 - Present | 2757 | 1280 |
| Web of Science Core Collection* | Web of Knowledge | 1975 - Present | 2108 | 612 |
| CINAHL | EBSCO | 1982 - Present | 2381 | 1861 |
| Total |  |  | 8953 | 5455 |

\*Science Citation Index Expanded (1975-present) ; Social Sciences Citation Index (1975-present) ; Arts & Humanities Citation Index (1975-present) ; Conference Proceedings Citation Index- Science (1990-present) ; Conference Proceedings Citation Index- Social Science & Humanities (1990-present) ; Emerging Sources Citation Index (2005-present)
No other database limits were used than those specified in the search strategies

### Medline

(exp Back Pain / OR exp Neck Pain / OR exp Osteoarthritis / OR (backache OR back-ache OR back-pain* OR neck-pain* OR osteoarthrit* OR spondylosis*).ab,ti,kw.) AND (Nomograms / OR ((*Models, Statistical /) AND (*Prognosis/ OR *Predictive Value of Tests/ OR *Probability/ OR *Diagnosis/ OR *Risk Assessment/ OR *Outcome Assessment, Health Care/ OR * Patient Outcome Assessment)) OR ((*Models, Statistical /) AND (Prognosis/ OR Predictive Value of Tests/ OR Probability / OR Diagnosis/ OR Risk Assessment/ OR Outcome Assessment, Health Care/ OR Patient Outcome Assessment) AND (Validation Study/ OR Quality Control/ OR “Sensitivity and Specificity”/)) OR ((diagnos* OR prognostic* OR predict* OR probab* OR risk*) AND (model* OR tool OR tools OR score*)).ti. OR ((diagnos* OR prognostic* OR predict* OR probab* OR risk*) ADJ3 (model* OR tool OR tools OR score*) ADJ10 (validat* OR develop* OR compar* OR test* OR evaluat* OR creat* OR accura* OR sensitiv* OR specific* OR perform* OR discriminat* OR calibrat* OR update* OR optimi* OR qualit*)).ab.) NOT (news OR congres* OR abstract* OR book* OR chapter* OR dissertation abstract*).pt.

### Embase

(backache/exp OR ‘neck pain’/de OR osteoarthritis/exp OR (backache OR back-ache OR back-pain* OR neck-pain* OR osteoarthrit* OR spondylosis*):ab,ti) AND (‘diagnostic model’/de OR ‘prognostic model’/de OR ‘predictive model’/de OR ((model/mj OR ‘disease model’/mj OR ‘population model’/mj OR ‘ scoring system’/mj OR nomogram/mj) AND (prognosis/mj OR prediction/mj OR probability/mj OR diagnosis/mj OR ‘diagnostic accuracy’/mj OR ‘risk assessment’/mj OR ‘outcome assessment’/mj)) OR ((model/de OR ‘disease model’/de OR ‘population model’/de OR ‘ scoring system’/de OR nomogram/de) AND (prognosis/de OR prediction/de OR probability/de OR diagnosis/de OR ‘diagnostic accuracy’/de OR ‘risk assessment’/de OR ‘outcome assessment’/de) AND (‘validation process’/de OR ‘validation study’/de OR ‘quality control’/de OR ‘sensitivity and specificity’/de OR ‘predictive value’/de)) OR ((diagnos* OR prognostic* OR predict* OR probab* OR risk*) AND (model* OR tool OR tools OR score*)):ti OR ((diagnos* OR prognostic* OR predict* OR probab* OR risk*) NEAR/3 (model* OR tool OR tools OR score*) NEAR/10 (validat* OR develop* OR compar* OR test* OR evaluat* OR creat* OR accura* OR sensitiv* OR specific* OR perform* OR discriminat* OR calibrat* OR update* OR optimi* OR qualit*)):ab) NOT ([conference abstract]/lim)

### Web of science

TS=((backache OR back-ache OR back-pain* OR neck-pain* OR osteoarthrit* OR spondylosis*)) AND (TI=((diagnos* OR prognostic* OR predict* OR probab* OR risk*) AND (model* OR tool OR tools OR score*)) OR TS=((diagnos* OR prognostic* OR predict* OR probab* OR risk*) NEAR/2 (model* OR tool OR tools OR score*) NEAR/10 (validat* OR develop* OR compar* OR test* OR evaluat* OR creat* OR accura* OR sensitiv* OR specific* OR perform* OR discriminat* OR calibrat* OR update* OR optimi* OR qualit*))) NOT DT=(Meeting Abstract OR Meeting Summary)

### CINAHL

(MH Back Pain + OR MH Neck Pain + OR MH Osteoarthritis + OR TI(backache OR back-ache OR back-pain* OR neck-pain* OR osteoarthrit* OR spondylosis*) OR AB(backache OR back-ache OR back-pain* OR neck-pain* OR osteoarthrit* OR spondylosis*)) AND (((MM Models, Statistical) AND (MM Prognosis OR MM Predictive Value of Tests OR MM Probability OR MM Diagnosis OR MM Risk Assessment OR MM “Outcomes (Health Care)” OR MM Outcome Assessment)) OR ((MH Models, Statistical) AND (MH Prognosis OR MH Predictive Value of Tests OR MH Probability OR MH Diagnosis OR MH Risk Assessment OR MH “Outcomes (Health Care)” OR MH Outcome Assessment) AND (MH Validation Studies OR MH Quality of Health Care OR MH “Sensitivity and Specificity”)) OR (TI(diagnos* OR prognostic* OR predict* OR probab* OR risk*) AND (model* OR tool OR tools OR score*)) OR AB((diagnos* OR prognostic* OR predict* OR probab* OR risk*) N2 (model* OR tool OR tools OR score*) N10 (validat* OR develop* OR compar* OR test* OR evaluat* OR creat* OR accura* OR sensitiv* OR specific* OR perform* OR discriminat* OR calibrat* OR update* OR optimi* OR qualit*)).ab.)

